# The relationship between ethnicity and socioeconomic deprivation as determinants of health: a systematic review

**DOI:** 10.1101/2024.03.06.24303819

**Authors:** Melanie Coates, Aroon Bhardwaj Shah, Richard Igwe, Yize I Wan

## Abstract

**Background:** Ethnicity and socioeconomic status (SES) are well known social determinants of health. However, the impact of the intersection between ethnicity and SES on health remains poorly understood, with many studies examining these factors separately.

**Methods:** We conducted a systematic review using MEDLINE (PubMed), EMBASE, and The Cochrane Library databases. Studies were eligible if they compared outcomes defined by mortality, attendance, readmission, or hospital length of stay, for any acute or chronic illness, according to one or more measures of both ethnicity and SES in adult patients (≥18 years of age) attending primary care or admitted to hospital.

**Results:** Nine studies met eligibility criteria. There was significant heterogeneity in cohort demographics, key variables, and outcome measures. Therefore, qualitative analysis was used. Definitions and categories of ethnicity were inconsistent, using race, country of origin, and sociocultural characteristics. Definitions of SES varied, with studies using between one and seven metrics. Different sub-categories were often used, even when the same metric was utilised. Primary outcomes were mortality (n=7) or admissions-related (n=2). Approaches varied between studies, regarding cause, time periods, and end points. Of those looking at mortality, four reported seeing an interaction between ethnicity and SES groups on outcomes.

**Conclusions:** Heterogeneity in the categorisation of ethnicity and SES is a barrier to research and understanding of health inequalities. This could be tackled by standardising data collection in healthcare routine data nationally and internationally, to enable translation of information between settings. For SES, using multifaceted methods could better capture the complexity of this factor.

## Introduction

Social determinants of health are a significant public health challenge, and a major driver of health inequity worldwide. Defined by the WHO as the conditions in which people are born, grow, live, work and age, they encompass the non-medical factors that shape health.[1] Two of the major determinants of health inequity are socioeconomic status (SES) and race and/or ethnicity. Historically, the term ethnicity referred more to a person’s cultural identity, and race to ancestral origin and physical characteristics.[2] It is now widely recognised that the terms race and ethnicity are social constructs, with evolving definitions and differing usage between regions.[3] Ethnicity and race are multifaceted terms which may encompass ancestry, culture, identity, language, religion and/or physical appearance, amongst other factors. Given the varied use of these terms, we use the term ethnicity throughout to refer to both race and ethnicity.

There is a well-established link between ethnicity and SES with health separately. In all countries, regardless of income level, health outcomes follow a social gradient: the lower the socioeconomic position, the worse the health status.[4] Low SES is associated with increased mortality around the globe, including in the UK, Europe, US, Australia, and Sub-Saharan Africa.[5–8] In many countries, low SES and minority ethnic background are associated with more acute hospital utilisation, including emergency department visits and emergency hospital admissions.[9–12] For people of minoritised ethnic groups, those with long term conditions have worse health than their White counterparts.[13,14] In the US between 2000-2017, adults of Non-Hispanic Black ethnicity had the highest mortality,[15] while in Canada, those from minoritised ethnic groups had lower hospital utilisation and cancer screening uptake.[16] In Brazil, hospitalisation in multimorbid patients is greater for those of lower SES and Black ethnicity, compared to less deprived and White patients.[17]

It is widely acknowledged that SES and ethnicity are connected. However, the relationship between ethnicity and SES is complex, with many minoritised ethnic groups experiencing poorer SES. For example, in the USA, the poverty rate is higher in all non-White ethnic groups compared to White.[18] In the UK, people of Black, Asian, and minority ethnic groups are 2.5 times more likely to be in relative poverty, and 2.2 times more likely to be in deep poverty than White people.[19] The COVID-19 pandemic has once again highlighted these inequalities, with socially disadvantaged and minoritised ethnic groups disproportionately affected.[20,21]

Despite this, the impact of the intersection of ethnicity and SES together on health remains poorly understood. There is a lack of research examining their interaction in determining health outcomes, with many studies only examining these risk factors separately. Research in this area is further complicated by differing definitions and measurements of ethnicity and SES.[22] Therefore, this systematic review aims to describe current knowledge on the interaction between ethnicity and socioeconomic deprivation in patients requiring primary and secondary healthcare services. We aim to summarise the characteristics of studies, highlight the evidence for interaction and synthesise findings and gaps in knowledge for future work.

## Methods

This systematic review was conducted in accordance with the Preferred Reporting Items for Systematic Reviews and Meta-Analyses (PRISMA) reporting guidelines.[23] The study protocol was pre-registered with the International Prospective Register of Systematic Reviews (PROSPERO; CRD42021257352).[24]

### Study selection

A study was eligible if it compared outcomes according to one or more measures of both ethnicity and socioeconomic deprivation in adult patients (≥18 years of age) attending primary care or admitted to hospital. Outcomes were defined as: 1) mortality, 2) repeated attendance or readmission, and 3) hospital length of stay, for any acute or chronic illness requiring access to healthcare services. We also examined retention in care as an additional outcome measure. Prospective and retrospective studies were included. Case reports and case series, as well as non-research publications such as literature reviews, editorials and correspondences were excluded. Only full text articles in English were included, to permit maximum data extraction.

### Search strategy

Three databases were searched from inception through 17 October 2022: MEDLINE (PubMed), Excerpta Medica dataBASE (EMBASE), and The Cochrane Library. Different combinations of keywords related to (a) ethnicity or race, (b) socioeconomic status, (c) health care use or mortality, and (d) association or relationship were used. The full search strategy is presented in Supplementary Materials.

### Data extraction

Data extraction was performed independently by four independent investigators (YW, MC, AB, and RI) using predefined data extraction forms. A random check of 10% of the cases was done at the inclusion and extraction staged to ensure concordance. Extracted data included study characteristics (country, year, number of centres, total sample size, inclusion criteria, exclusion criteria, definition of ethnicity, categorisation of ethnicity, definition of SES, and categorisation of SES), patient characteristics (age, sex, ethnicity, SES), and outcome data (mortality, hospital admission or readmission, or hospital length of stay). Where definition of ethnicity was not reported in the study itself, definitions were taken from the original source or database, where available.

### Quality of evidence and risk of bias

Quality of evidence and individual study risk of bias within studies was assessed using the National Institutes of Health (NIH) quality assessment tools. Assessment was done at the study level and in outcome reporting. Poor quality studies were excluded from data synthesis.

### Data synthesis

Quantitative analysis was anticipated with plan to conduct a meta-analysis of effect sizes with random effects. The following criteria was predefined to be met for data synthesis: 1) minimum of 5 studies; and 2) consistency in study population, definition of exposures and outcomes, predefined subgroup characteristics of disease and time period. Due to insufficiently homogenous study populations, studies were critically appraised to summarise the best available evidence though qualitative data synthesis. Qualitative data synthesis was focused on descriptive statistics and aimed to report on range and heterogeneity in key variables.

## Results

### Search results and study characteristics

A total of 2223 unique records were identified. 2159 records were excluded following title and abstract screening. The full text was reviewed for 64 records, following which a further 55 were excluded (Figure 1). A total of nine records met inclusion criteria (Table 1).[25–33] Most of these studies (n=7) were conducted in North America (six in the USA; one in Canada), with two in Europe (one in the UK; one in Belgium) (Figure 2a). There was significant heterogeneity between studies due to variations in cohort demographics, definitions of key variables, and outcome measures. Given the insufficient methodological homogeneity between studies, quantitative analysis was deemed inappropriate, therefore studies were analysed using qualitative assessments. In NIH quality assessment, seven studies were determined to be fair and two were good (Figure 2b).

**Figure 1.**
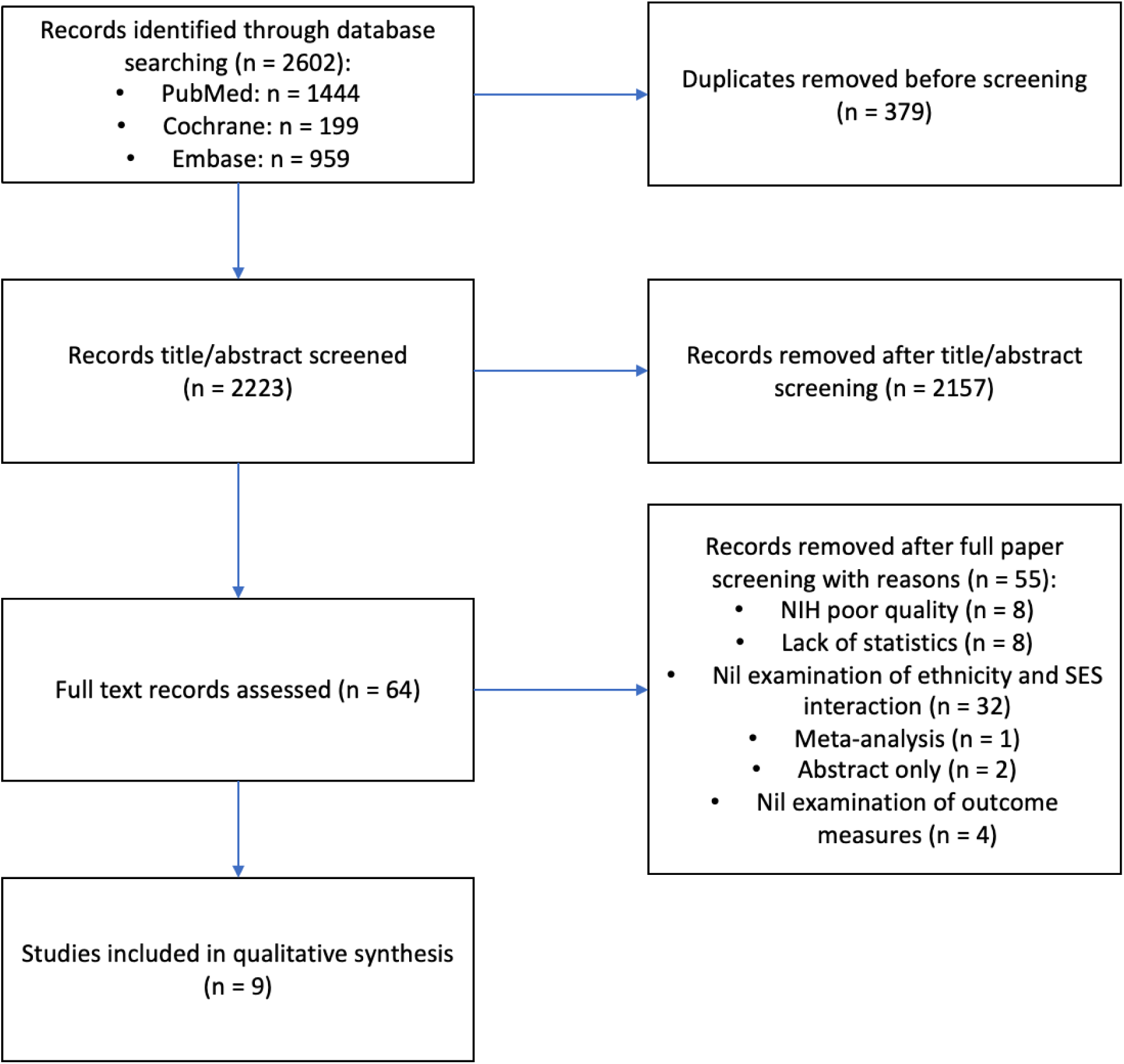
PRISMA flowchart.

**Figure 2.**
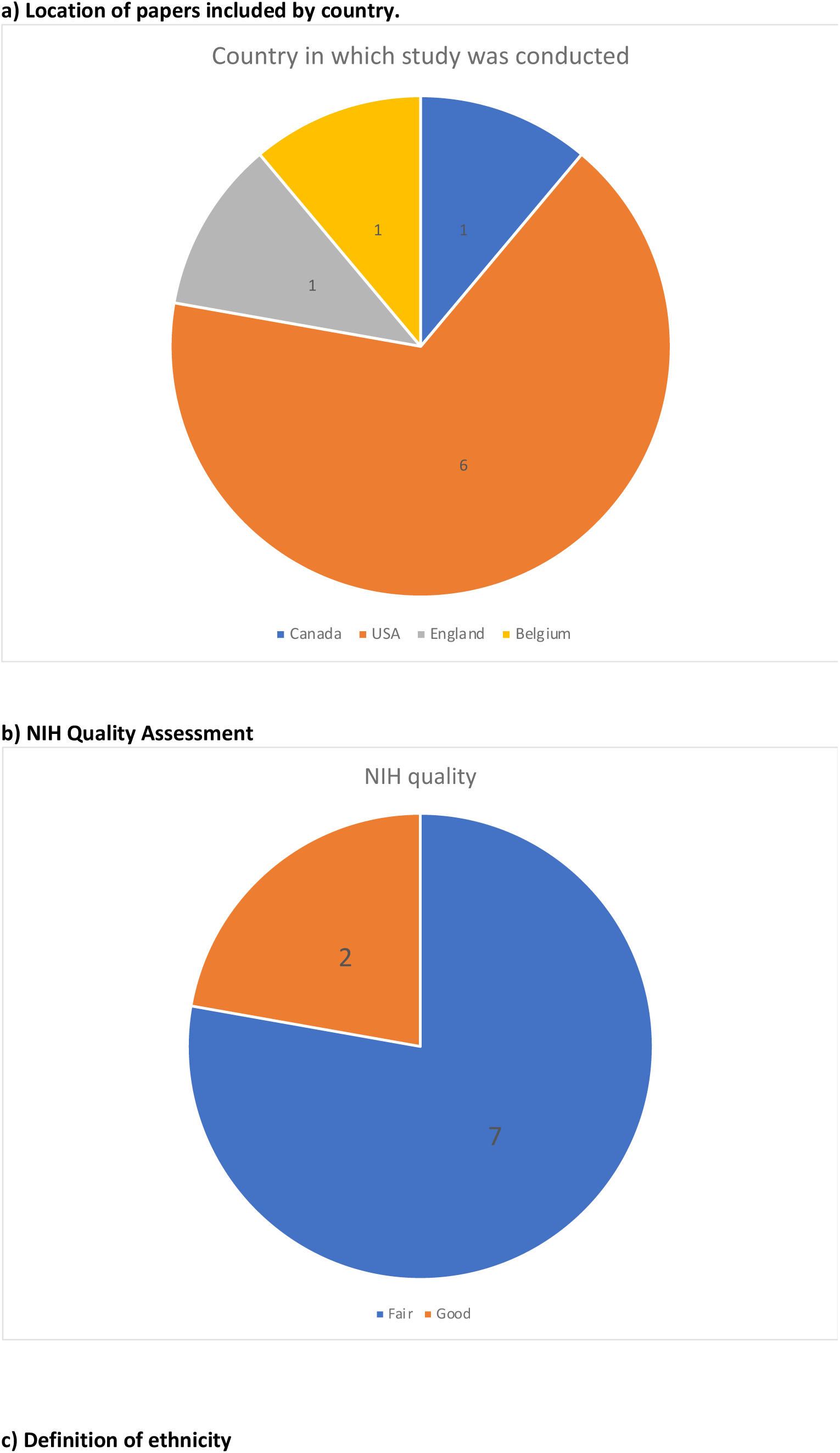
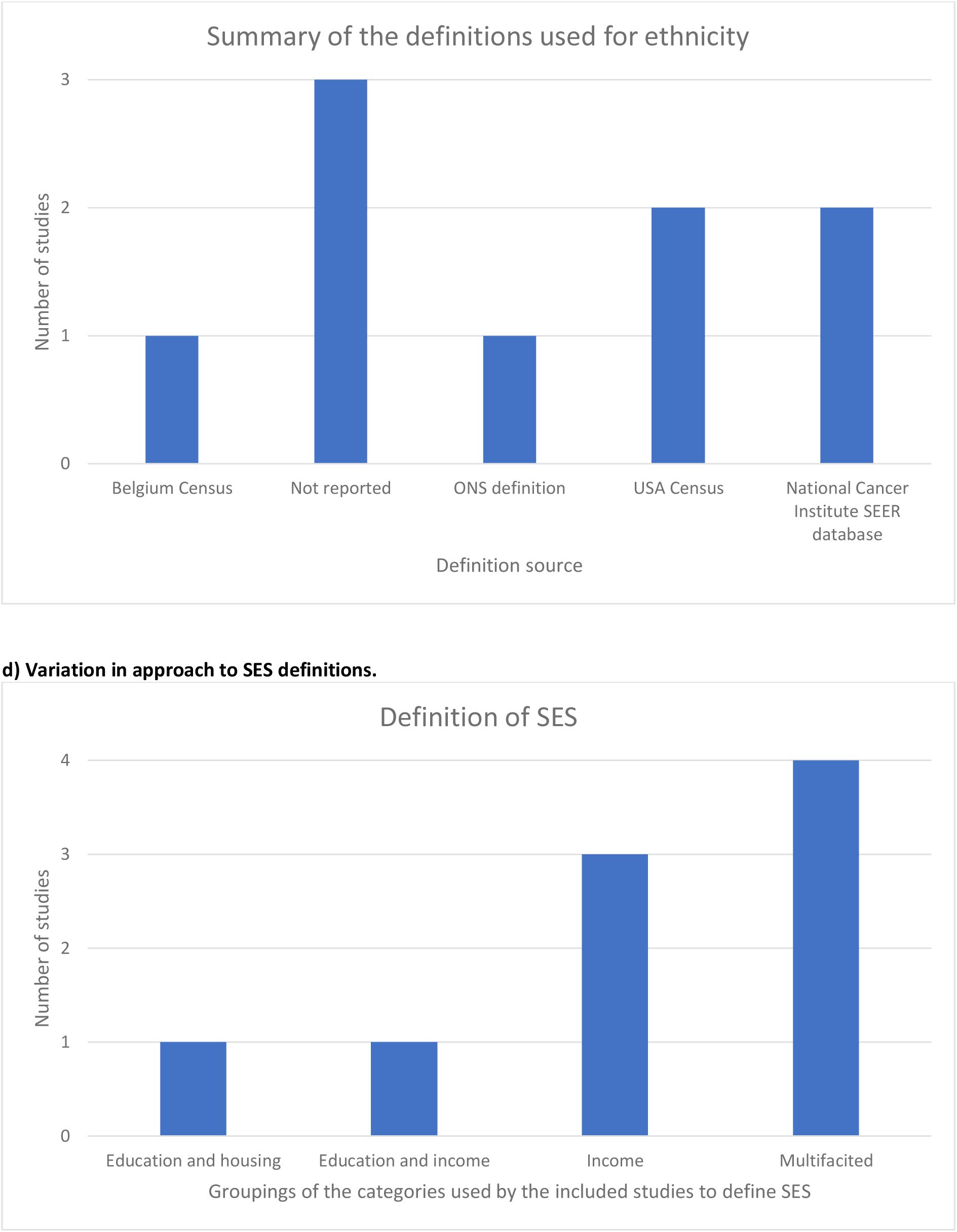
Qualitative assessment of included studies.

**Table 1.** Summary table of included studies. Studies listed by first author and year. SES: socioeconomic status, HR: hazard ratio, OR: odds ratio, CVD: cardiovascular disease

| Study | Country | Total n | Number of categories - ethnicity | Categories used for ethnicity | SES – metrics used | Number of categories - SES | Primary outcome category | Primary Outcome | Effect measure | Adjusted | Key findings | Study design | NIH quality |
| --- | --- | --- | --- | --- | --- | --- | --- | --- | --- | --- | --- | --- | --- |
| Kim C, 2005[25] | USA | 207625 | 2 | Black, White | Education and income | 2 | Mortality | Cause-specific mortality due to breast cancer (vs. no reported death or death due to other causes) | OR | Yes | Less than high school education was associated with decreased hospital mortality from CVD in both Black and White women. However, less than a high school education was associated with lower breast cancer mortality among White women but not among Black women. Similar ORs were seen for self-reported income as for education level. | Ecological | Fair |
| Du XL, 2007[26] | USA | 18492 | 3 | African American (Non-Hispanic Black), Caucasian (Non-Hispanic White), Other | Multifaceted (Researcher defined: education, income, and poverty) | 4 | Mortality | All-cause mortality (patients with stage II or III colon-cancer); colon-cancer specific mortality | HR | Yes | Increased risk of mortality from colon cancer for those of African American ethnicity. This was explained substantially by differences in SES, with a larger proportion (70%) of African American patients with colon cancer in the poorest quartiles of SES compared with Caucasians (21%). | Cohort | Fair |
| Freeman V, 2011[27] | USA | 833 | 2 | African American, Non-Hispanic White | Multifaceted (modified Browning and Cagney formula) | 4 | Mortality | Cause-specific mortality due to prostate cancer | HR | No | African Americans were significantly more likely to die of prostate cancer relative to non-Hispanic Whites, which disappeared when accounting for concentrated disadvantage. | Cohort | Fair |
| Tian N, 2011[28] | USA | 44515 | 3 | African American, Hispanic, Non-Hispanic White | Poverty | 3 | Mortality | Cause-specific mortality (breast cancer) | OR | Yes | Low and middle SES were more likely to show significant racial disparities of breast cancer late-stage diagnosis and mortality rates, for African American and Hispanic women when compared to non-Hispanic White women. | Ecological | Fair |
| Vandenheede H, 2011[29] | Belgium | 299837 | 2 | Belgian, North African | Education and housing status | 6 | Mortality | Cause-specific mortality (diabetes-related) | Mortality Rate Ratio | Yes | North Africans have a higher diabetes-related mortality compared to Belgians. These differences in diabetes-related mortality largely disappear when differences in education are taken into account. | Ecological | Fair |
| Aseltine R, 2015[30] | USA | 63445 | 5 | Asian, Black, Hispanic, Other, White | Income | N/A | Admission | Readmission (for all causes) to the same hospital within 30 days of discharge (patients initially hospitalised with chest pain or heart failure) | OR | Yes | Hispanic patients were more likely to be readmitted within 30 days of discharge following hospitalisation for heart failure compared to White patients, both before and after adjusting for SES, patient sickness, and patient's insurance status. Those of Black ethnicity were significantly more likely to be readmitted within 30 days of discharge following hospitalisation for chest pain, compared White ethnicity (although the difference was not seen after controlling for SES, comorbidities, and insurance status). | Ecological | Fair |
| Nishino Y, 2015[31] | England | 445504 | 6 | Asian, Black, Mixed, Other, South Asian, White | Multifaceted (Index of Multiple Deprivation (IMD)) | 5 | Admission | Inpatient admissions and readmissions due to diabetes | RR | No | South Asian ethnicity was associated with higher admission risk, but lower re-admission risk when compared with White ethnicity. Readmission risk increased with IMD among White British but not other ethnicities. There was a statistically significant interaction between ethnicity and deprivation. | Cohort | Good |
| Abdel-Rahman O, 2019 | Canada | 80121 | 5 | American Indian, Asian, Black Non-Hispanic, Hispanic, White Non-Hispanic | Multifaceted (Yost et al. and Yu et al.) | 3 | Mortality | Colon cancer-specific survival (defined as the time from colon cancer diagnosis until death from colon cancer) | HR | Yes | Higher SES was associated with better survival for White, Black, and Hispanic ethnicities. No significant change in survival rates was found when comparing the different ethnic groups at different SES levels | Ecological | Good |
| Rotsides J, 2020[33] | USA | 45940 | 5 | Asian, Black, Hispanic, Other, White | Income | 4 | Mortality | Overall survival | HR | Yes | Black ethnicity (compared to White) and low SES are associated with worse survival. Survival for those of Black ethnicity within the lowest SES group was lower than White ethnicity within the highest SES group. When controlling for HPV status, this disparity remained significant. | Cohort | Fair |

### Definition and categorisation of ethnicity

Five studies referred to both race and ethnicity.[26–28,30,33] Two solely discussed race[25,32] and two solely discussed ethnicity.[29,31] The two studies only using ethnicity were both by authors in Europe (UK and Belgium).

Methods for defining ethnicity varied between studies (Figure 2c). Three studies did not report a definition of ethnicity.[28,30,33] These studies used pre-existing databases as the source of ethnicity data: one used the National Cancer Database (NCDB),[33] one used the Texas Cancer Registry and the Vital Statistic Unit database,[28] and one used the Acute Care Hospital Inpatient Discharge Database (HIDD), where ethnicity was self or observer reported. Two studies used the National Cancer Institute (NCI) SEER (Surveillance, Epidemiology, and End Results) database,[26,32] which defines ethnicity using race and country of origin. This database distinguishes between race and ethnicity, separating Hispanic from non-Hispanic ethnic groups. Race and ethnicity are reported in five mutually exclusive categories: Non-Hispanic White, Non-Hispanic Black, Non-Hispanic Asian/Pacific Islander (API), Non-Hispanic American Indian/Alaska Native (AI/AN), Hispanic (all races).

US Census data was used by two studies, which uses a social definition of race to encompass racial or national origin or sociocultural groups.[25,27] One study used the UK Hospital Episode Statistics (HES) database, which uses the UK Office for National Statistics (ONS) definition of ethnic group,[31] which defines ethnicity as based on their culture, family background, identity, or physical appearance. One study used nationality either currently or at birth, with data taken from the Belgium Census.[29] Categorisation of ethnicity was inconsistent between studies. Studies used between two to six categories of ethnicity, with the most frequent number being two categories (n=3).[25,27,29] Methods of categorising ethnicity were not reported in three studies.[28,30,33] The most frequently used categorisations were using Census categories by three studies: USA[25], UK ONS[31] and Belgian Census[29]. The next most frequent was the NCI SEER categorisation used by two studies.[26,32] Studies from the USA used eight categories of ethnicity in total (White, Black, African-American, Asian, Hispanic, Non-Hispanic White, Non-Hispanic Black, and Other).[25–28,30,33] The study from Canada used four categories: White Non-Hispanic, Black Non-Hispanic, Asian, and Hispanic.[32] In the UK study, six categories were used: White, Mixed, South Asian, Black, Asian, Other.[31] In the study from Belgium, only two categories were used (Belgian or North African).[29] North African ethnicity was defined as nationality, either current or at birth, as being from one of several countries in the North of Africa.

### Definition and categorisation of socioeconomic status

Several different methods for defining SES were taken by the studies investigated, with studies using varying numbers of metrics to define SES (Figure 2d). For the purposes of this analysis those that used three or more metrics have been classified as having taken a multifaceted approach. Four studies used a multifaceted approach. Of these, three used existing methods of defining and categorising SES. Abdel-Rahman et al. used a composite score by Yost et al. and Yu et al.,[32] which uses seven parameters (unemployment percent, working-class percent, percent below 150% of the poverty line, education index, median house value and median household income). Freeman et al. used a modified version of the Browning and Cagney formula for census tract-level concentrated disadvantage score, defined by four parameters: % in poverty + % unemployed + % female-headed households + (100 - % college graduate).[27] The study by Nishino et al. used Index of Multiple Deprivation (IMD), which is determined using seven variables: income, employment, health, education, housing services, crime, and the living environment.[31] The remaining study, by Du et al., defined SES with a researcher-determined composite score using education (the percentage of adults aged >25 years who had <12 years of education), median annual household income, and percentage of people living below the poverty line.[26]

Two studies used a combination of two metrics to define SES. Of these, one used education and income (Kim et al.).[25] These metrics were chosen as other measures of SES were not predictive of mortality after adjusting for education and income in previous analyses. The other study used education and housing status (Vandenheede et al.).[29] Two studies used income as a single parameter to define SES (Aseltine et al. and Rotisides et al.),[30,33] and one used percentage of population under the poverty level (Tian et al.).[28] The three studies that used a single metric were all situated in the USA.

The categorisation of SES also varied between studies, with studies using between two and five categories for SES. Of those that took a multifaceted approach, one study used tertiles,[32] two used quartiles,[26,27] and one used quintiles.[31] Of the study using education and income, education was defined by three categories: less than high school, high school (with and without a degree), and some college or more.[25] Self-reported income was used in secondary analyses. In the study that used education and housing status, education was categorised in line with the International Standard Classification of Education (ISCED) (five categories: pre-primary, primary, lower secondary, upper secondary and tertiary education).[29] Housing status was based on tenure status as well as housing quality and categorised into 6 categories: low-, mid- and high-quality tenants and low-, mid- and high-quality owners. Of the studies using income as a proxy for SES, one did not report which categories were used,[30] and one used four categories: median household income over $63000, $48000-$62999, $38000-$47999, or under $38000. The study using poverty used three categories: <10%, 10-20% and >20% population living below the poverty level.[28]

### Outcomes

Primary outcomes chosen by the studies examined were mortality or admissions related. The most frequently used outcome was mortality (n=7). Five studies examined cause-specific mortality, one looked at all-cause mortality, and one looked at both all-cause and cause-specific mortality.

Of those looking at cause-specific mortality, two studies examined breast cancer-specific mortality.[25,28] Kim *et al.* included all deaths recorded via death certificate between 1979 and 1989,[25] and Tian *et al.* looked at breast cancer cases between 1995 to 2005.[28] One study (Abdel-Rahman *et al.*) examined colon-cancer specific survival (defined as the time from colon cancer diagnosis until death from colon cancer).[32] One study (Freeman *et al.*) looked at death from prostate cancer as the underlying cause, in patients diagnosed between 1^st^ January 1986 and 31^st^ December 1990, who were followed up until 31^st^ December 2006. Death certificates were obtained for all known decedents, or cause of death determined through the National Death Index.[27] One study (Vandenheede *et al.*) examined diabetes-related mortality (defined as death certificates with diabetes as an underlying cause of death and with diabetes as one of the causes of death) according to the mortality register data between 2001 and 2005.[29]

One study looked at all-cause mortality (Rotsides *et al.*). The primary end point was 3 and 5-year overall survival in patients with oropharyngeal squamous cell carcinoma between 2010 to 2016.[33] One study looked at both all-cause and colon-cancer specific mortality (Du *et al.*). Survival was calculated in months from the date of diagnosis to the date of death or to the date of last follow-up (up to 11 years).[26]

Two studies examined hospital admission: one for first admission or readmission due to diabetes, defined as a hospital admission with primary or secondary diagnosis from the ICD 10 codes E10—E14 (Nishino *et al.*),[31] and one on readmission (for all causes) to the same hospital within 30 days of discharge (Aseltine *et al.*).[30]

### Relationship between ethnicity and SES

Four studies reported that ethnic disparities were seen, and that they were likely due to SES through adjusted covariate modelling, with one study reporting that SES plays only a small role on ethnic disparities. The remaining studies described differences seen in outcomes between racial and SES groups through stratification.

Of the two studies examining outcomes defined by admission, Nishino *et al.* reported that readmission risk increased with increasing deprivation by SES among White British but no other ethnicities.[31] Aseltine *et al.* reported that, compared to those of White ethnicity, patients of Hispanic ethnicity were significantly more likely to be readmitted within 30 days of discharge following hospitalisation for heart failure, both before and after adjustment for SES, severity of illness, and insurance status (Medicare, Medicaid, Other Payer *vs* Private Payer).[30] Those of Black ethnicity were significantly more likely to be readmitted within 30 days of discharge following hospitalisation for chest pain, compared to those of White ethnicity, although the difference was not seen after controlling for patient socioeconomic status, comorbidities, and insurance status.

Of the studies that examined mortality, four described an interaction between ethnicity and SES groups on outcomes. Abdel-Rahman *et al.* reported that lower SES index was associated with worse survival in the four racial groups studied (White Non-Hispanic, Black Non-Hispanic, Asian, and Hispanic).[32] Kim at el. found that lower than high school level education was associated with decreased hospital mortality from cardiovascular disease in both Black and White women. However, lower than high school level education was associated with lower breast cancer mortality among White women but not among Black women. Similar ORs were seen for self-reported income as for education level.[25] Rotsides *et al.* found that survival for those of Black ethnicity within the lowest SES group was lower than White ethnicity within the highest SES group.[33] Tian *et al.* found that low and middle SES groups were more likely to show significant ethnic disparities of breast cancer mortality rates, for African American and Hispanic women when compared to Non-Hispanic White women.[28]

Of the studies with mortality as an outcome, three attempted to understand the cause for ethnic disparities, finding they were likely due to SES. Du *et al.* found there was an increased risk of mortality from colon cancer for those of African American ethnicity.[26] This was explained substantially by differences in SES with a larger proportion (70%) of African American patients with colon cancer in the poorest quartiles of SES compared with Caucasian patients (21%). Freeman *et al.* reported that patients of African American ethnicity were significantly more likely to die of prostate cancer relative to Non-Hispanic White patients. This difference disappeared when accounting for concentrated disadvantage.[27] Vandenheede *et al.* found that people from North Africa had higher diabetes-related mortality compared to Belgian people. These differences in diabetes-related mortality largely disappeared when differences in education were considered.[29]

## Discussion

This systematic review highlights the challenges faced in understanding the complex relationship between ethnicity, SES, and health outcomes. Despite an extensive literature search and broad inclusion criteria, only nine studies were identified as specifically aimed to understand the relationship between ethnicity and SES as the primary study objective. Most studies supported pre-existing observational data and the hypothesis that worsening SES increases disparities in health outcomes experienced by minoritised ethnic groups. However, a major barrier we found in interpreting the data is the heterogeneity in defining and categorising ethnicity and SES, limiting generalisability and transferability of results. What’s more, none of the studies we identified built on prior knowledge and did not reference the other studies we examined.

Despite studies into the interaction of ethnicity and SES, there was no standardised way of reporting or analysing data, preventing effective comparison of studies. We found significant heterogeneity in reporting of ethnicity with variations in definition, categorisation, and data collection. These variations may be due to geographical differences in demographics, understanding of ethnicity, different historical and cultural contexts, and political or social factors. For example, in the USA, use of the ethnic group Hispanic is common, however it was not used in other countries and regions. As there are larger numbers of people immigrating between areas of Latin America and the USA, this ethnic group may be more represented in data collection categories. Whereas in Europe, this category is not seen as commonly in data collection, likely due to lower levels of immigration from these areas and thus smaller populations who may identify with this group. In different regions, understandings of ethnicity may vary, therefore we would expect variations in how people self-identify and report. In areas with higher levels of migration and subsequent multiple generations, this may also impact how people self-identify, due to the environment they were raised in versus the ancestral origins. As ethnicity is not a fixed concept, interpretations may also shift over time due to changes in societal norms.

Some papers used only a single measure of SES, such as income. As SES is a complex concept, a single variable may not capture the nuances of SES to fully understand an individual’s social status and economic situation. Therefore, by taking more factors into account and standardising these measures, we can capture different dimensions of SES to more accurately to depict someone’s SES and reduce bias. Therefore, multifaceted reporting of SES may enable a more comprehensive and accurate representation of an individual’s SES. Although most papers included in this review used a multifaceted reporting of SES, included measures and components varied widely, limiting comparability.

Heterogeneity in classification of ethnicity and SES presents an obstacle for data collection, analysis, and reporting, not only limited to academic research. Ethnic and racial inequality data is often used as a proxy measure for social processes of racialisation such as experiences of racism, social practices (such as diet), and genetic traits (such as thalassemia) within health and when addressing risk factors.[34] Classifications of ethnicity and SES inform public health monitoring systems, which subsequently are used to identify inequalities and monitor trends. This has implications for policy making and public health practice, such as in resource allocation and activating responses. Therefore, how we classify groups is crucial in capturing meaningful data for public health research and practice.[35]

Ethnicity is a complex socio-political construct that encompasses genetic make-up, shared origin, language, and cultural traditions, therefore there is no universally accepted definition of the term and reaching consensus on categorisation is difficult.[34,36] In some cases, race, ethnicity, and nationality may be used interchangeably, or with considerable overlap, and there is often variation within and between countries. For example, we found in Belgium, North African nationality is equated with ethnicity. In North America in particular, the terminology race is often used. Wide overarching groups are often used, which do not represent the heterogeneity of regions. What’s more, inconsistencies are seen between countries, due to differing racial and ethnic make-up of countries and the subjective nature of ethnicity. For example, in the USA often Hispanic and Latino are included, a category not often seen in Europe, likely reflecting differing demographics and immigration patterns.[37] These discrepancies are even notable within countries, for example in the UK, Census data is collected differently in each region. For example, England and Wales use three categories for Black ethnicity (Black African, Black Caribbean, Other Black), whereas Northern Ireland uses two (Black African or Black Other).[38,39] Comparatively, Scotland uses six categories for White (Scottish, Other British, Irish, Gypsy/Traveller, Polish, Other White), whereas England and Wales use five (English/Welsh/Scottish/Northern Irish/British, Irish, Gypsy or Irish Traveller, Roma and Other).[38,40] Some countries do not collect ethnicity data, such as in France and Germany. These differences are further complicated by the fact that changing demographics due to immigration means historical categories may quickly become outdated and insufficient to capture current or future populations.[41] therefore, fixed-response categories may be inadequate to capture the complexities of ethnicity as an identity, as it encompasses a wide range of experiences and backgrounds and is often context-dependent in nature.

SES is a similarly complex construct, comprising diverse socioeconomic factors such as economic resources, social and work status, power, and prestige.[42,43] it is often measured by using different variables of income, education, and occupation. Often studies use only one measure of SES, may use few categories, and/or measure at a single time point. As variables are not interchangeable and may capture different aspects of health, this may not be sufficient to capture the complexity of SES and may act to obscure social gradients and full understanding of the impact of SES.[44] What’s more, SES may affect health differently at different times during the life course, or impact at different levels (individual, household, community etc).[43,45] Differing ways of defining and measuring SES between countries poses additional challenges when comparing internationally. For example, in the USA income is often used, whereas the UK often uses Area Deprivation Index.

By over assuming homogeneity of groups, there is a risk of population profiling when estimating public health risk, for example biological reductionism when considering ethnicity. Using broad categories, such as Black, Asian, or White, may prevent identification of variations in sub-groups and mask inequalities. Furthermore, ethnicity and SES may further interact with other SDOH, to cause differential health effects. Therefore, it is important not to consider groups as homogenous populations, and to bear in mind other influential factors and the complex intersection of factors when considering health outcomes and inequalities.

The studies identified in this review did not report on difficulties in data collection or defining ethnicity or SES. There are some efforts to standardise defining and categorising ethnicity and SES within countries.[46,47] However, it is widely known however that there remains little to no standardisation between countries. Collection of data on ethnicity and SES through standardised categories in routine healthcare use would be of benefit locally, nationally, and internationally, to enable understanding of inequalities and comparison between regions. Although we recognise there are many challenges in standardising SES and ethnicity categorisation, improving international consensus would support transferability of research findings within countries and internationally. Using multiple categories and definitions would better capture the complexities of defining ethnicity and SES. It is also important to consider how data is collected, as this will affect findings, for example when comparing self-reported vs clinician reported information. More research is needed to reach consensus on the optimal way to categorise these populations, and the best proxies to use. This would benefit health research, to improve data collection and standardise comparison within countries and internationally.

### Strengths and limitations

Strengths of this work include that it was preregistered with an *a priori* protocol. To our knowledge, no other systematic reviews have examined this topic. To be as extensive as feasible we searched three separate search engines without time limits, through conception until recent. However, limits of this work were the unsuitability for quantitative analysis. Therefore, we have aimed to describe as much as possible to improve current knowledge through qualitative analysis. There remains potential for publication bias. Due to the breadth of our search terms, variations in the structure of study reporting, and exclusions of non-English articles, it is possible that some studies were missed.

## Conclusions

To date, most health and health care research has focused on the effects of ethnicity and SES separately, making it difficult to understand the complex interactions and cumulative effects of these factors on health and health inequalities. Heterogeneity in the categorisation of ethnicity and SES both within and between countries is a barrier to research and understanding of health inequalities. This could be tackled by standardising data collection in healthcare routine data nationally and internationally, to enable translation of information between settings. For SES, using multifaceted methods can better capture the complexity of this factor. For ethnicity, self-identification, using comprehensive ethnicity categories with multiple options and being sensitive to geographical and regional differences will be of benefit. More research is needed to investigate the role of ethnicity and SES together on health. Having a thorough understanding of these relationships will enable us to understand the effect on populations with different or multiple marginalising experiences, and to identify health inequalities. This will enable us to identify groups in need, target customised interventions and work towards eliminating health inequities.

## Declarations

### Conflicts of interest/Competing interests

All authors declare no competing interests related to the submitted work.

### Funding

The authors declare that no funds, grants, or other support were received during the preparation of this manuscript.

### Availability of data and material

The data that support the findings of this study are openly available. The datasets used and/or analysed during the study are available from the corresponding author upon request.

### Code availability

Not applicable.

### Authors’ contributions

MC and YIW wrote the study protocol and analysis plan. All authors reviewed and approved the final manuscript. YIW was responsible for the study concept and study design. All authors were involved in study identification and data extraction. Analyses were undertaken by AB, MC and YIW. MC, AB, and YIW wrote the manuscript.

### Ethics approval

Ethical approval was not required.

### Consent to participate

Not applicable.

### Consent for publication

Not applicable

## Abbreviations

EMBASE: Excerpta Medica dataBASE
HES: Hospital Episode Statistics
HIDD: Hospital Inpatient Discharge Database
IMD: Index of Multiple Deprivation
NCDB: National Cancer Database
NCI: National Cancer Institute
ONS: Office for National Statistics
PRISMA: Preferred Reporting Items for Systematic Reviews and Meta-Analyses
SEER: Surveillance, Epidemiology, and End Results
SES: socioeconomic status

## Notes

### Competing Interest Statement

The authors have declared no competing interest.

