## Supplementary material for "The relationship between ethnicity and socioeconomic deprivation as determinants of health: a systematic review"

### Supplementary Material 1 – Search Strategies

#### Search strategy – MEDLINE & Embase

Database: Embase <1974 to 2022 October 17>, Ovid MEDLINE(R) ALL <1946 to October 17, 2022>

Search Strategy:

--------------------------------------------------------------------------------

1	("ethnicit*" or "ethnic group").ti,ab.

2	("race" or "racial group*").ti,ab.

3	exp "Ethnic groups"/

4	exp "Continental Population Groups"/ or Racial Groups/

5	("deprivation" or "socioeconomic group" or "socioeconomic factor" or "socioeconomic status" or "socio-economic group").ti,ab.

6	("socio-economic factors" or "socio-economic status" or "wealth" or "poverty").ti,ab.

7	("predict*" or "associat*" or "relat*").ti.

8	("primary care attendance" or "primary care utilisation" or "primary care consultation" or "primary care appointment" or "general practice attendance").ti,ab.

9	("general practice utilisation" or "general practice consultation" or "general practice appointment").ti,ab.

10	exp Primary Health Care/

11	exp General Practice/

12	("hospital admission" or "patient admission" or "hospitalization" or "hospitalisation" or "secondary care admission").ti,ab.

13	exp Hospitalization/

14	exp "Appointments and Schedules"/

15	("mortality" or "hospital readmission" or "patient readmission").ti,ab.

16	exp Mortality/

17	(1 or 2 or 3 or 4) and (5 or 6) and 7 and (8 or 9 or 10 or 11 or 12 or 13 or 14 or 15 or 16)

18	17 use medall

19	17 use oemezd

20	20 or 21

21	remove duplicates from 22

22	limit 23 to english language

23	limit 24 to humans

24	limit 25 to human

#### Search strategy - Cochrane

#1	("ethnicity"):ti,ab,kw OR ("ethnic group"):ti,ab,kw OR ("ethnic background"):ti,ab,kw OR ("ethnicities"):ti,ab,kw (Word variations have been searched)

#2	("race"):ti,ab,kw OR ("racial group"):ti,ab,kw (Word variations have been searched)

#3	MeSH descriptor: [Ethnic Groups] explode all trees

#4	MeSH descriptor: [Continental Population Groups] explode all trees

#5	("deprivation"):ti,ab,kw OR ("socioeconomic group"):ti,ab,kw OR ("socioeconomic factor"):ti,ab,kw OR ("socioeconomic status"):ti,ab,kw OR ("socio-economic group"):ti,ab,kw (Word variations have been searched)

#6	("socio-economic factors"):ti,ab,kw OR ("socio-economic status"):ti,ab,kw OR ("wealth"):ti,ab,kw OR ("poverty"):ti,ab,kw (Word variations have been searched)

#7	("predict*"):ti,ab,kw OR ("associat*"):ti,ab,kw OR ("relat*"):ti,ab,kw (Word variations have been searched)

#8	("primary care attendance"):ti,ab,kw OR ("primary care utilisation"):ti,ab,kw OR ("primary care consultation"):ti,ab,kw OR ("primary care appointment"):ti,ab,kw OR ("general practice attendance"):ti,ab,kw (Word variations have been searched)

#9	("general practice utilisation"):ti,ab,kw OR ("general practice consultation"):ti,ab,kw OR ("general practice appointment"):ti,ab,kw (Word variations have been searched)

#10	MeSH descriptor: [Primary Health Care] explode all trees

#11	MeSH descriptor: [General Practice] explode all trees

#12	("hospital admission"):ti,ab,kw OR ("patient admission"):ti,ab,kw OR ("hospitalization"):ti,ab,kw OR ("hospitalisation"):ti,ab,kw OR ("secondary care admission"):ti,ab,kw (Word variations have been searched)

#13	MeSH descriptor: [Hospitalization] explode all trees

#14	MeSH descriptor: [Appointments and Schedules] explode all trees

#15	("mortality"):ti,ab,kw OR ("hospital readmission"):ti,ab,kw OR ("patient readmission"):ti,ab,kw (Word variations have been searched)

#16	MeSH descriptor: [Mortality] explode all trees

#17	(#1 OR #2 OR #3 OR #4) AND (#5 OR #6) AND (#7) AND (#8 OR #9 OR #10 OR #11 OR #12 OR #13 OR #14 OR #15 OR #16)

### Supplementary Material 2 – Data Extraction Tables

#### Table 1. Study Characteristics

| **Title** | **First author** | **Year of publication** | **Country** | **Number of centres** | **Total n** | **Age in years: median (IQR) +/- mean (SD)** | **Male: n (%)** | **Patient inclusion criteria** | **Patient exclusion criteria** | **Method** | **Study design** | **Statistical analysis** | **NIH quality assessment** |
| --- | --- | --- | --- | --- | --- | --- | --- | --- | --- | --- | --- | --- | --- |
| Is education associated with mortality for breast cancer and cardiovascular disease among black and white women?[1] | Kim C | 2005 | USA | Not reported (National database) | 207625 | Overall not reported | 0 | Black women and non-Hispanic white women aged ≥20 years | Women with missing data on education or income level; women of other racial or ethnic groups (e.g., Hispanic, Asian, Native American) because of limited numbers and to reduce errors introduced by multiple comparisons | Statistical modelling | Ecological | Association modelling | Fair |
| Racial disparities and socioeconomic status in association with survival in a large population-based cohort of elderly patients with colon cancer[2] | Du XL | 2007 | USA | Not reported (National database) | 18,492 | Median age at diagnosis was 77 years for Caucasians, 75 years for African Americans, and 76 years for others | 40.7%, 36.3%, 46.8% | Diagnosed with stage II or III colon cancer at age ≥65 years between 1992 and 1999 that was their first and only primary tumor | Patients who did not have full coverage of both Medicare Parts A and B or who were members of a Health Maintenance Organization in the year when their diagnosis was made to ensure the completeness of Medicare claims | Statistical modelling | Cohort | Association modelling | Fair |
| Association of census tract-level socioeconomic status with disparities in prostate cancer-specific survival.[3] | Freeman V | 2011 | USA | 4 | 833 | Mean 8.6 (6.9) | 833 | African-American and non-Hispanic white men diagnosed with adenocarcinoma of the prostate between January 1, 1986 and December 31, 1990 | T1a-stage lesions which are considered clinically insignificant; incomplete data; residential address at diagnosis could not be matched to its corresponding census tract geocode | Statistical modelling | Cohort | Association modelling | Fair |
| Spatial association of racial/ethnic disparities between late-stage diagnosis and mortality for female breast cancer: where to intervene?[4] | Tian N | 2011 | USA | Not reported (Texas State database) | 44,515 | Not reported | 0 | Individual breast cancer incidence and mortality cases from 1995 to 2005 | Not reported | Statistical modelling | Ecological | Descriptive | Fair |
| Ethnic differences in diabetes-related mortality in the Brussels-Capital Region (2001-05): the role of socioeconomic position.[5] | Vandenh-eede H | 2011 | Belgium | Not reported (BrusselsCapital Region) | 299837 | 25-74 years all | Not reported | Belgian and North African men and women aged 25-74 living in the BCR at the time of the census. | Not reported | Stratification | Ecological | Descriptive | Fair |
| Connecticut Hospital Readmissions Related to Chest Pain and Heart Failure: Differences by Race, Ethnicity, and Payer[6] | Aseltine R | 2015 | USA | 30 | CP = 23,450, HF = 39,995 | Mean age chest pain = 62.1, mean age heart failure = 77.3 | CP = 11066, HF = 10011 | Adult patients 18 years of age or older with an index admission for Chest Pain (DRG 313) or Heart Failure and Shock (DRG 291 and 292) during the 2008- 2012 CMS fiscal years | Not reported | Logistic models for clustered data using generalized estimating equations (GEE) in R | Ecological | Association modelling | Fair |
| Inequality in diabetes-related hospital admissions in England by socioeconomic deprivation and ethnicity: facility-based cross-sectional analysis.[7] | Nishino Y | 2015 | England | 355 | 5147859 (445504 diabetic admissions) | Not reported | Not reported | Non-obstetric patients over 16 years old in all NHS facilities in England | Outside England; referral cases; obstetric cases; patients <16 years old; data from facilities with less than 80% diagnostic record coverage; data from facilities with less than 80% ethnicity data coverage | Statistical modelling | Cohort | Association modelling | Good |
| Outcomes of non-metastatic colon cancer patients in relationship to socioeconomic status: an analysis of SEER census tract-level socioeconomic database[8] | Abdel-Rahman O | 2019 | Canada | Not reported (National database) | 80121 | Not reported | 39489 (49.3%) | Colon adenocarcinoma diagnosis; stages I, II, III (according to American Joint Committee on Cancer 7th staging sys- tem); complete information about SES group at the time of diagnosis; diagnosis year between 2004-2015; upfront treatment with an oncologic surgical resection (including resection of the primary and regional lymph node dissection). | Cases with rectal primary or non-adenocarcinoma histology | Statistical modelling | Ecological | Association modelling | Good |
| Socioeconomic and Racial Disparities and Survival of Human Papillomavirus-Associated Oropharyngeal Squamous Cell Carcinoma[9] | Rotsides J | 2020 | USA | Not reported (National database) | 45940 | 60 (20-90) | 38038 (82.8%) | Patients with oropharyngeal squamous cell carcinoma diagnosed between 2010-2016 | Unknown HPV status; under 18; T0 tumours; unknown outcomes; death within 6 months of diagnosis; prior malignancies; lack of treatment/unknown treatment | Cox regression analyses | Cohort | Association modelling | Fair |

#### Table 2. Ethnicity and SES Metrics

| **Study** | **Use of ethnicity-related terms** | **Ethnicity - source of data** | **Ethnicity - definition reported in study** | **Ethnicity - categorisation method** | **Ethnicity - number of categories used** | **Ethnicity - names of categories** | **Ethnicity - n in each category** | **SES - metrics used** | **SES - number of categories** | **SES - names of categories** | **SES - n in each category** |
| --- | --- | --- | --- | --- | --- | --- | --- | --- | --- | --- | --- |
| Kim C, 2005[1] | Race | US Census Bureau National Longitudinal Mortality Study database (NLMS) | Not reported | US Census | 2 | Black, White | Black 21303, White 186322 | Education and income | 2 | Education: less than high school, high school, some college or more ed self-reported annual income: (<$10,000, $]0,000-$19,999, or ≥$20,000) | Black: less than high school = 9367, high school = 7202, some college or more = 4734 White: less than HS 50558, HS 78795, some college or more = 56969 |
| Du XL, 2007[2] | Race and ethnicity | National Cancer Institute SEER database (Medicare linked database) | Not reported | National Cancer Institute SEER database | 3 | African American (Non-Hispanic Black), Caucasian (Non-Hispanic White), Other ethnicities (which were combined because of small numbers) | African American (Non-Hispanic Black) 1320, Caucasian (Non-Hispanic White) 15913, Other 1259 | Multifaceted (Researcher defined: education, income, and poverty) | 4 | Quartiles 1 highest to 4 lowest | First quartile 4281+73+270, Second quartile 4226+122+271, Third quartile 4140+206+282, Fourth quartile 3266+919+436 |
| Freeman V, 2011[3] | Race and ethnicity | Census USA 1990 | Not reported | Census USA 1990 (only looked at African American and Non-Hispanic White groups) | 2 | African American, Non-Hispanic White | African American 320, Non-Hispanic White 513 | Multifaceted (modified Browning and Cagney formula) | 4 | 1st quartile, 2nd quartile, 3rd quartile, 4th quartile | Not reported |
| Tian N, 2011[4] | Race and ethnicity | Texas Cancer Registry and the Vital Statistic Unit, Texas Department of State Health Services | Not reported | Not reported | 3 | African American, Hispanic, Non-Hispanic White | Not reported | Poverty | 3 | Low (>20% population living below the poverty level), middle (10% ~20%) and high SES group (<10%) | Not reported |
| Vandenheede H, 2011[5] | Ethnicity (ethnic origin) | Belgian census (2001) | North African ethnicity is defined as being of North African origin. If either current nationality or nationality of birth is Algerian, Egyptian, Libyan, Moroccan or Tunisian, people are then considered as being a part of the North African community. | Belgian census (2001) | 2 | Belgian, North African | Not reported | Education and housing status | 5 | Education: pre-primary, primary, lower secondary, upper secondary and tertiary. Housing: low-, mid- and high-quality tenants and low-, mid- and high-quality owners. | Not reported |
| Aseltine R, 2015[6] | Race and ethnicity | Self reported or observer reported (Acute Care Hospital Inpatient Discharge Database (HIDD)) | Not reported | Not reported | 5 | Asian, Black, Hispanic, Other, White | Chest pain: Asian 126, Black 3622, Hispanic 2664, Other 680, White 16385 Heart failure: Asian 133, Black 4639, Hispanic 2328, Other 828, White 32057 | Income | N/A | Not reported | Not reported |
| Nishino Y, 2015[7] | Ethnicity | UK Hospital Episode Statistics (HES) data | Not reported | ONS Census | 6 | Asian, Black, Mixed, Other, South Asian, White | Not reported | Multifaceted (Index of Multiple Deprivation (IMD)) | 5 | 1 (Least deprived), 2, 3, 4, 5 (most deprived) | Not reported |
| Abdel-Rahman O, 2019[8] | Race | National Cancer Institute SEER database (Census Tract‐Level SES Database) | Not reported | National Cancer Institute SEER Database | 5 | American Indian, Asian, Black Non-Hispanic, Hispanic, White Non-Hispanic | American Indian 285 (0.4%), Asian-Pacific Islander 398 (0.5%), Black non-Hispanic 8751 (10.9%), Hispanic 6748 (8.4%), White non-Hispanic 8442(10.5%) | Multifaceted (Yost et al. and Yu et al.) | 3 | Group 1, group 2, group 3 | Group 1 24368, group 2 27838, group 3 27915 |
| Rotsides J, 2020[9] | Race and ethnicity | National Cancer Database (NCDB) | Not reported | Not reported | 5 | Asian, Black, Hispanic, Other, White | Asian 497 (1.1%), Black 3226 (7%), Hispanic 1473 (3.2%), Other 588 (1.3%), White 40156 (87.4%) | Income | 4 | Over $63000, $48000-$62999, $38000-$47999, Under $38000 | Over $63000 - 16151 (35.2%), $48000-$62999 - 12436 (27.1%), $38000-$47999 - 10172 (22.1%), Under $38000 - 7070 (15.4%) |

#### Table 3. Primary Outcomes

| **Study** | **Primary Outcome** | **Effect measure 1** | **Effect size 1 (95% CI), unadjusted** | **p value 1, unadjusted** | **Adjusted?** | **Adjustment variables** | **Effect size 1 (95% CI), adjusted** | **p value 1, adjusted** |
| --- | --- | --- | --- | --- | --- | --- | --- | --- |
| Kim C, 2005[1] | Cause-specific mortality due to breast cancer (vs no reported death or death due to other causes) | Odds ratio | Less than a high school education: white women OR 1.5 (1.2-1.9); Black women OR 1.8 (0.9-3.6) | Not reported | Yes | Age (years), marital status (married/not married), 16 and urban or rural residence | Less than a high school education was associated with lower breast cancer mortality among White women (OR, 0.73; 95% CI, 0.6-0.9) but not among Black women (OR, 1.1; 95% CI, 0.5-2.3) |  |
| Du XL, 2007[2] | All-cause mortality (patients with stage II or III colon-cancer); colon-cancer specific mortality | Hazards ratio | African American 1.18 (1.10-1.27), Other 0.97 (0.89-1.06) | Not reported | Yes | Age, sex, comorbidity, tumour stage, grade, year of diagnosis, Surveillance Epidemiology and End Results region, optimal therapy, and radiation and treatment | 1.10 (1.02‚Äì1.19) African-American, 0.94 (0.86‚Äì1.03) others | not reported |
| Freeman V, 2011[3] | Cause-specific mortality due to prostate cancer | Hazards ratio | 0.96 (0.83-1.11) | 0.61 | No | N/A | N/A | N/A |
| Tian N, 2011[4] | Cause-specific mortality (breast cancer); late-stage diagnosis | Odds ratio | African American 33.76 (CI: 23.96-47.57), Hispanic 30.39 (CI: 22.09-41.82) | <0.000001 | Yes | SES | African American 18.9 (CI: 12.79-26.44), Hispanic 11.64 (CI: 8.29-16.34) | <0.000001 |
| Vandenheede H, 2011[5] | Cause-specific mortality (diabetes-related) | MRR (mortality rate ratio) | North African 1.62 (95% CI 1.11‚Äì2.37) in men, 3.35 (95% CI 2.08‚Äì5.41) women | N/A | Yes | Housing + education | 1.34 (0.95-1.92) for men and 1.85 (1.09-3.12) for women |  |
| Aseltine R, 2015[6] | Readmission (for all causes) to the same hospital within 30 days of discharge (patients initially hospitalised with chest pain or heart failure) | Odds ratio | Asian 0.73 (0.34-1.56), Black 1.19 (1.04-1.37), Hispanic 1.07(0.91-1.25), Other 0.63 (0.45-0.90) | Asian 0.417, Black 0.012, Hispanic 0.44, Other 0.012 | Yes | Length of stay, patient comorbidities, patient SES, and patient insurance (Medicare, Medicaid, Other Payer vs Private Payer) | Asian 0.38 (0.38-1.83), Black 0.93 (0.80-1.07), Hispanic 0.79 (0.66-0.94), Other 0.64 (0.45-0.91) | Asian 0.644, Black 0.302, Hispanic 0.007, Other 0.013 |
| Nishino Y, 2015[7] | Inpatient admissions and readmissions due to diabetes | Risk ratio | IMD 1 Least deprived (RR - 1), IMD 2 (RR1.16 [1.13-1.19]), IMD 3 (RR 1.35 [1.32-1.39]), IMD 4 (RR 1.68 [1.63-1.73]), IMD 5 Most deprived (RR 2.08 [2.0-ì2.14]) | p ≥ 0.001 for all | No | N/A | N/A | N/A |
| Abdel-Rahman O, 2019[8] | Colon cancer-specific survival (defined as the time from colon cancer diagnosis till death from colon cancer) | Hazards ratio | Not reported | Not reported | Yes | Age, sex, race, and side | Group 1 vs group 3: 1.257; 1.190-1.328 | p<0.001 |
| Rotsides J, 2020[9] | Overall survival | Hazards ratio | Black 1.51 (1.42-1.59) Hispanic 1.09 (0.92-1.28) Asian 1.14 (0.88-1.48) | Black <0.01, Hispanic 0.33, Asian 0.32 | Yes | Age, sex, comorbidity, HPV status, insurance type, income level, tumour stage, and treatment | Black (1.22 1.11-1.34 p<0.01) Hispanic (0.88 0.74-1.04 p-0.12) Asian (1.08 0.83-1.4 p-0.58) | Black P < 0.01  Hispanic P – 0.12  Asian P 0.58 |

**Table 4. Study characteristics - other outcomes**

| **Study** | **Outcome 2** | **Effect measure 2** | **Effect size 2 (95% CI), unadjusted** | **p value 2, unadjusted** | **Adjustment variables** | **Effect size 2 (95% CI), adjusted** | **p value 2, adjusted** | **Outcome 3** | **Effect measure 3** | **Effect size 3 (95% CI), unadjusted** |
| --- | --- | --- | --- | --- | --- | --- | --- | --- | --- | --- |
| Kim C, 2005[1] | CVD death (vs no reported death or death due to other causes) | Odds ratio | Less than a high school education was associated with a greater likelihood of CVD mortality among both Black (odds ratio [OR], 6.9; 95% CI, 4.2-11.5) and White women (OR, 4.8; 95% CI, 4.4-5.4) compared with some college education or more | Not reported | Age, marital status, and urban or rural residence | Less education was still associated with greater CVD mortality among Black women (OR, 1.8; 95% CI, 1.03-3.0) and White women (OR, 1.4; 95% CI, 1.3-1.6). |  | Self-reported income |  | General patterns of effects reported in the table for education level were similar to the effects for self-reported income (results not shown) |
| Du XL, 2007[2] | 10-year colon-cancer specific mortality | Hazards ratio | 1.26 (1.11-1.43) African American, 1.17 (1.01-1.35) Others | Not reported, adjusted for age; sex; comorbidity; tumour stage; grade; year of diagnosis; Surveillance, Epidemiology, and End Results region; optimal therapy; and radiation and treatment |  | 1.16 (1.01-1.33) African American, 1.12 (0.97-1.30) Others | Not reported | N/A | N/A | N/A |
| Freeman V, 2011[3] | N/A | N/A | N/A | N/A | N/A | N/A | N/A | N/A | N/A | N/A |
| Tian N, 2011[4] | N/A | N/A | N/A | N/A | N/A | N/A | N/A | N/A | N/A | N/A |
| Vandenheede H, 2011[5] | N/A | N/A | N/A | N/A | N/A | N/A | N/A | N/A | N/A | N/A |
| Aseltine R, 2015[6] | Likelihood of readmission (for all causes) to the same hospital within 30 days of discharge (compared to White patients) - heart failure | Odds ratio | Asian 0.98 (0.62-1.57), Black 1.03 (0.94-1.14), Hispanic 1.30 (1.15-1.47), Other 0.80 (0.65-0.99) | Asian 0.939, Black 0.491, Hispanic 0.000, Other 0.037 | Length of stay, patient comorbidities, patient SES, and patient insurance (Medicare, Medicaid, Other Payer vs Private Payer) | Asian 1.00 (0.62-1.60), Black 1.02 (0.92-1.12), Hispanic 1.26 (1.11-1.43), Other 0.81 (0.66-1.00) | Asian 0.985, Black 0.730, Hispanic 0.000, Other 0.048 | N/A | N/A | N/A |
| Nishino Y, 2015[7] | Multiple regression model of relationship between hospital admission due to diabetes (ethnicity) | Risk ratio | White (RR 1), Asian ( 1.02 [0.87-1.21]), Black (1.22 [1.09-1.37]), Mixed (RR 1.49 [1.29-1.72]), Others (1.32 [1.21-1.43]), South Asian (2.62 [2.51-2.74]) | Asian (0.783), Black (0.001), Mixed (<0.001), Others (>0.001), South Asian (< 0.001) | N/A | N/A | N/A | Multiple regression model of relationship between hospital admission due to diabetes (IMD Ethnicity interaction) | Risk ratio | IMD 2 : (Mixed - 0.83 [0.68-1.01]) (South Asian 0.91 [0.86-0.97]) (Black 1.23 [1.07-1.42]) (Asian 1.22 [0.95-1.57]) (Others 1.03 [0.92-1.116]), IMD 3: (Mixed 1.15 [0.97-1.37]),(South Asian (0.85 [ 0.81‚Äì0.91]), (Black 1.2 [1.05-1.36]), (Asian 0.83 [0.65-1.06]), IMD 4 (Mixed 0.73 [0.61-0.86])(South Asian 0.81 [0.77-0.86]) (Black 1.12 [0.99-1.26])(Asian 1.1 [0.89-1.36]). IMD 5 (Mixed 0.86 [0.73-1.01])(South Asian 0.81 [0.77-0.85]) (Black 1.10 [0.97-1.24]) (Asian 1.07 [0.87-1.32]) |
| Abdel-Rahman O, 2019[8] | Impact of SES index on colon cancer, specific survival in specific subgroups (according to colon cancer side and stage, and according to patients’ sex and race) | Hazards ratio | White Non-Hispanic Group 3 1.220 (1.144-1.301), Group 1 1.166 (1.099-1.237).  Black non-Hispanic Group 3 1.366 (1.122-1.664), Group 1 1.224 (0.985-1.520)  Hispanic - Group 3 1.283 (1.076-1.531), Group 1 1.175 (0.975-1.416).  Asian/Pacific Islander - Group 3 1.543 (1.249-1.907), Group 1 1.059 (1.308-1.087) | N/A |  |  |  |  |  |  |
| Rotsides J, 2020[9] | Overall Mortality Mean Household income (<$38000 as reference) | Hazards ratio | $38000-47999 (0.71 0.65-0.77 p<0.01), $48000-62999 (0.58 0.53-0.63 p<0.01), $63000+ (0.44 0.4-0.58 p<0.01) | $38000-47999  (p<0.010  $48000-62999 (p<0.01)  $63000+ (p<0.01) | Age, sex, comorbidity, HPV status, insurance type, ethnicity, tumour stage, and treatment | $38000-47999 (0.85 0.78-0.93 <0.01) $48000-62999 (0.75 0.69-0.81 p-0.01) $63000+ (0.63 0.58-0.69 p-0.01) | $38000-47999(p<0.01)  $48000-62999 (p<0.01)  $63000 (p<0.01) |  |  |  |
